# Age at separation, residential mobility, and depressive symptoms among twins in late adolescence and young adulthood: a FinnTwin12 cohort study

**DOI:** 10.1101/2022.07.05.22277258

**Authors:** Zhiyang Wang, Alyce Whipp, Marja Heinonen-Guzejev, Jaakko Kaprio

## Abstract

**Objectives:** Leaving the parental home at an early age represents a major life event for an adolescent and may predispose them to poor mental health. This study aims to examine the effect of age at separation and residential mobility (from residential records) on depressive symptoms among twins in late adolescence and young adulthood. Age at separation of twins in a twin pair was used as a proxy indicator for the age of leaving the parental home

**Methods:** The participants (n=3055) are from the FinnTwin12 cohort. Residential mobility consisted of the number and total distance of moves before age 17. First, we used linear regression to assess the association of age at separation and residential mobility with log-transformed General Behavior Inventory (GBI) scores at age 17 and in young adulthood. Then, the mixed model for repeated measures (MMRM) was used to visualize the GBI scores’ trajectory and test the associations, controlling for “baseline” state.

**Results:** In the adjusted linear regression, compared to twins separated before age 17, twins who separated in the later three groups all were significantly lower on log-transformed GBI scores at age 17. The two groups of twins who separated after age 19.5 scored lower GBIs in young adulthood. Compared to twins who never moved before age 17, moving twice led to a lower GBI score. In MMRM, twins who separated later all had lower GBI scores in young adulthood. Twins who moved twice or more had lower GBI scores in young adulthood.

**Conclusion:** The current study provides valid evidence of the family influence on depressive symptoms in later adolescence and young adulthood. A strong association between residential mobility and depressive symptoms was affirmed, although further research is still needed in the future.

## 1. Introduction

The global burden of major depressive disorder (MDD) is getting progressively worse since the last century [1]. High-income areas had a higher prevalence of MDD [2]. In 2019, according to the Global Burden of Diseases, the age-standardized prevalence of depressive disorder was 5626.6 per 100,000 people in Western Europe and was higher than the global level [3]. Data for the general US population shows the prevalence of depression in adolescents and young adults has been increasing since 2005 [4], [5]. Multiple risk factors for depression in childhood and adolescence have been identified such as poor education, stressful life events, and family predisposition [6].

During childhood, moving to a new location exposes children to greater social network loss than adults and their parents are less able to give them the attention they need. Moving to a new location is regarded as a type of negative life experience that is associated with later depression [7], [8]. Residential instability or mobility describes those children who have an unstable life and are more frequently exposed to this deleterious experience. Numerous studies have suggested that high residential instability leads to a larger risk of many psychological or psychiatric outcomes, including depression [7], [9]–[11]. However, the measure of residential mobility is often limited by recall bias from children’s or their parents’ questionnaires [9]. Longitudinal residential records from the national system could quantify the cumulative effect of mobility to enrich the assessment of the strength of this relationship [11].

In late adolescence and young adulthood, people will gradually move out of their parental homes due to various reasons. Anxiety driven by early separation from parents has been connected to a series of negative mental conditions such as complicated grief, more severe symptoms of depression, or panic disorder at a later time [12]–[14]. Parental support acts like a “safety net” to enhance the resilience in coping with challenges and to sustain the successful transition to adulthood of the child [15], [16]. Family closeness, support, attentiveness, and other positive factors are associated with a later timing of leaving, while teenagers living with step-parents or incomplete families tend to leave early due to a lack of resources, a trend seen in multiple countries [17]–[19].

Leaving home also means parting with siblings. Because of the similar age and same generation, siblings spend more time together and have more mutual influence. Positive sibling bonding is capable of relieving detrimental parental influence [20]. For a relatively extreme example in China, AIDS orphans who were separated from their siblings suffered a higher level of anxiety, depression, anger, and dissociation [21]. As a special case of siblings, in general, twins have a closer relationship and reciprocity with each other than non-twin siblings [22], [23]. The separation between cotwins, alternatively termed “relational shift”, breaks the intense intimacy and dependency and can be accompanied by emotional turmoil and confusion due to individuation [24]. Separation is an important challenge for twins during childhood and adolescence.

The age at separation between cotwins reflects the length of time that twins stay together in the parental home, which is likely associated with the amount of family support, degree of mental maturity, and preparation time for twins to address the separation challenge, connecting to later mental health. On the other hand, we also assume the time of separation is when at least one of the cotwins leaves the parental home. Unlike questions about the family environment in surveys, age at separation by residential history in twins could be used as an objective measure to describe the separation time validly and reduce information bias. Therefore, in this study, we aim to determine: 1) the association between age at separation and depressive symptoms and the sex-specific effect, and 2) the association between residential mobility before age 17 and depressive symptoms, in late adolescence and young adulthood, based on the FinnTwin12 twin cohort.

## 2. Materials and methods

### 2.1 Participants

The study participants were from the FinnTwin12 cohort, which is a population-based prospective cohort among all Finnish twins born between 1983 and 1987. Participants were divided into two parts: intensive study and overall cohort. Overall 1035 families were invited to the intensive study from the overall (epidemiological) cohort (n=5184 twins at wave one) for psychiatric interviews, some biological samples, and additional questionnaires. A total of 1854 twins participated at age 14, and they were invited back for new interviews and other measurements as young adults. Interviews (n=1347) in young adulthood were completed for 73% of twins in the intensive study. All of twins in the overall cohort filled in the general age 17 questionnaire (wave three) and the non-intesive sample replied to young adult (wave four) questionnaires with 75% and 66% retention, respectively. The updated review of this cohort was published in 2019 [25].

The ethical committee of the Helsinki and Uusimaa University Hospital District and Indiana University’s Institutional Review Board approved the ethics of the study. All twins and their parents provided informed consent or written informed consent.

### 2.2 Measures

#### Age at separation

Age at separation was defined as the age that, for the first time, the twins no longer resided at the same address, with one or both twins moving to a new address as described elsewhere [26]. It was generated from the residential records linking to the Finnish Population information system since birth. The residential records included the information on the north and east coordinates (EUREF-FIN), and the age moving in and out of each address. Then, we categorized it into four groups: separated before age 17, between ages 17 and 19.5, between ages 19.5 and 22, and after age 22. Subsequentially, we created 2 variables for separation status (yes, no) at ages 17 and 22 based on the age at separation. Twins without valid information on age at separation (42 individuals) or twin pairs who never separated by the end of the follow-up in December 2020 (13 pairs) were excluded from the analysis with age at separation.

#### Residential mobility

Residential mobility consisted of two variables: the number of moves (never, once, twice, more than three times) and the total distance of moves (in kilometers) before age 17 accumulated across all moves. Both were generated from the residential records. If twins did not have address information due to missing addresses or living abroad for any period, their total distance of moves would be invalid and excluded from the corresponding analysis (122 individual twins). Overall, the mean total distance of moves before age 17 was 51.9 (SD 167.7) kilometers, and the total distance of moves was distributed in a right skewed fashion.

#### Depressive symptoms

We used the short-version General Behavior Inventory (GBI) to evaluate the depressive symptoms among twins [27]. It is a self-reported inventory designed to identify mood-related behaviors such as depressive and manic symptoms [28]. The short-scale version has 10 items and uses a 4-point Likert scale from 0 (never) to 3 (very often) to query the occurrence of depressive symptoms. The total score ranges from 0 to 30. This reliable measure has been used in many Finnish studies including FinnTwin12 [27], [29], [30]. To validate the GBI, we compared it to a DSM-IV diagnosis of MDD assessed by a structured interview (SSAGA) from the intenstive study [31]. In a logistic regression analysis, the GBI score in young adulthood strongly predicted MDD, with the area under the receiver operating characteristic curve (AUC) of 0.8321. The GBI was assessed at age 17 and in young adulthood. We also calculated the change of GBI score between the two stages. The mean age when twins provided the GBI assessment in young adulthood was 24.18 years (SD: 1.68), which was only adjusted in the analysis involving GBI in young adulthood.

Another two variables for the within-pair differences in GBI score were created at both stages. The variables were constructed by subtracting the GBI score of one cotwin from the score of the other cotwin, and then we took the absolute values. Higher values indicated a higher level of within-pair difference.

#### Covariates

There were seven covariates defined *a priori*: sex (male, female), zygosity (monozygotic (MZ), dizygotic (DZ), unknown), smoking (never, quit, occasional, current) in young adulthood, work status (full time, part-time, irregular, not working) in young adulthood, secondary level school (vocational, senior high school, none) in young adulthood, parental education, and age when twins provided the GBI assessment in young adulthood. If both parents of the twins had low education levels (less than high school, 12 year) or high education (12 or more years) levels, the parental education was classified as “limited” or “high”. If parents’ education levels were discordant, then it was classified as an intermediate level [32].

### 2.3 Statistical analysis

First, we excluded twins who were missing information on both depressive symptom assessments at ages 17 and in young adulthood, residential records, or any covariates. This left 3055 individual twins for analysis, of whom 2755 provided data at age 17 survey and 3029 in young adulthood. Because of the differential missing information pattern, sample sizes in different analyses were slightly different.

Due to the skewness of the GBI score at both ages, we added one to the GBI score and log-transformed it. Then, linear regression was used to examine the association of age at separation and residential mobility with log-transformed GBI scores at ages 17 and in young adulthood and change of GBI score. The pre-specified covariates were adjusted in the models. We used the univariate analysis of variance to determine the statistical differences between the mean GBI score for categorical covariates’ groups. Additionally, the separation before age 17 (no/no information/never lived together, yes) was further adjusted in the analysis with residential mobility

Further, since we had two measures of GBI score, the MMRM was chosen to visualize the trajectory of GBI score by age at separation for twins at age 17 and in young adulthood. This technique is able to consider the influence on the outcome (GBI in young adulthood) by both exposures of interest (fixed effect) and “baseline” GBI at age 17 (random effect) [33]. It also allowed us to handle the unevenness of missing information at different ages to increase the power. Covariates were adjusted for. Stratification analyses by sex were also performed, given that depression is more common in women than in men after puberty.

To evaluate the relationship between separation and change of twin similarity, at first, a bivariate cross-lagged path model was fitted with separation status and within-pair differences in GBI score at both stages with the maximum likelihood estimator (twin-pairs were analyzed as observations). This gauged the structural relationships of repeatedly measured variables. Two covariates were adjusted: sex and zygosity combination (male monozygotic (MMZ), female monozygotic (FMZ), male dizygotic (MDZ), female dizygotic (FDZ), and opposite-sex dizygotic (OSDZ)) and parental education. Twin pairs whose zygosity was unknown or who had missing information on separation status and within-pair differences in GBI score at both ages were excluded from the cross-lagged path model. Second, intraclass correlation coefficients (r) were calculated to determine the existence of a genetic variance component for depressive symptoms at both ages preliminarily, and we stratified the calculation by age at separation.

Additionally, we controlled for the cluster effect by twin pair in the regression analyses through “robust” standard errors. Regression coefficients and 95% confidence intervals (CIs) were reported. All statistical analyses were performed using Stata 17.0 (StataCorp, College Station, TX, USA) and Mplus 8.6 (Muthen & Muthen, Los Angeles, CA, USA).

## 3. Results

### 3.1 Demographic characteristics

A total of 3055 twins were included in the final analysis including 1279 twin pairs with both twins included and another 497 individual twins (i.e. their cotwin had not participated or otherwise excluded as defined above). As shown in Table 1, the mean score for GBI assessment at age 17 was 5.10, and in young adulthood was 4.42. The majority of twins were female (56.4%), dizygotic (60.7%), and reported never smoking in young adulthood (51.5%). For socioeconomic status, work status, secondary level school, and parental education all had significant between-group differences in GBI score at age 17 and young adulthood.

**Table 1:** Demographic, socioeconomic characteristics, and depressive symptoms for individual twins

| Characteristics | Individual n (%) | Mean (SD) or P-value |  |
| --- | --- | --- | --- |
|  |  | GBI at age 17 <sup>a</sup> | GBI in young adulthood <sup>b</sup> |
| **Overall** | 3055 | 5.10 (4.86) | 4.42 (4.68) |
| **Sex** |  | <0.0001 | <0.0001 |
| Male | 1331 (43.6) | 3.67 (3.90) | 3.62 (4.30) |
| Female | 1724 (56.4) | 6.15 (5.21) | 5.03 (4.87) |
| **Zygosity** |  | 0.00 | 0.06 |
| Monozygotic | 1059 (34.7) | 4.64 (4.61) | 4.14 (4.54) |
| Dizygotic | 1853 (60.7) | 5.32 (4.96) | 4.54 (4.68) |
| Unknown | 143 (4.7) | 5.71 (5.16) | 4.83 (5.58) |
| **Smoking** |  | 0.02 | 0.03 |
| Never | 1632 (53.4) | 4.43 (4.37) | 3.75 (4.08) |
| Quit | 340 (11.1) | 6.19 (5.44) | 4.68 (5.01) |
| Occasional | 308 (10.1) | 5.66 (5.01) | 4.47 (4.73) |
| Current | 775 (25.4) | 5.88 (5.32) | 5.68 (5.39) |
| **Work status** |  | 0.00 | 0.01 |
| Full-time | 1572 (51.5) | 4.92 (4.85) | 4.00 (4.28) |
| Part-time | 393 (12.9) | 5.06 (4.42) | 4.49 (4.32) |
| Irregular | 371 (12.1) | 5.08 (4.60) | 4.40 (4.35) |
| Not working | 719 (23.5) | 5.51 (5.21) | 5.30 (5.67) |
| **Secondary level school** |  | 0.02 | 0.03 |
| Vocational | 1035 (33.9) | 5.01 (4.76) | 4.42 (4.61) |
| Senior high school | 1843 (60.3) | 4.90 (4.58) | 4.10 (4.36) |
| None | 177 (5.8) | 7.87 (7.20) | 7.72 (6.72) |
| **Parental education** |  | 0.00 | 0.00 |
| Limited | 1758 (57.6) | 5.28 (4.96) | 4.58 (4.77) |
| Intermediate | 674 (22.1) | 4.88 (4.70) | 4.02 (4.26) |
| High | 623 (20.4) | 4.81 (4.72) | 4.37 (4.84) |

<sup>a</sup> 300 individual twins were missing because they did not provide GBI assessment at age 17
<sup>b</sup> 26 individual twins were missing because they did not provide GBI assessment in young adulthood

### 3.2 Association of age at separation with GBI

The sample sizes of each linear regression analysis are presented in Table 2. After adjustment for covariates, compared to twins separated before age 17, there were significant lower log-transformed GBI scores at age 17 in all three groups of twins, those who separated between ages 17 and19.5 (beta: -0.16, 95% CI: -0.30, -0.01), between ages 19.5 and 22 (beta: -0.23, 95% CI: -0.37, -0.08), and after age 22 (beta: -0.43, 95% CI: -0.59, -0.27). Moreover, twins who separated between ages 19.5 and 22 and after age 22 also had lower log-transformed GBI scores in young adulthood (beta: -0.18, 95% CI: -0.35, -0.00 and beta: -0.23, 95% CI: -0.42, -0.05, respectively).

**Table 2:** Association of age at separation and residential mobility with GBI using linear regression

| Characteristics | log-transformed GBI score at age 17 |  |  | log-transformed GBI score in young adulthood |  |  | Change of GBI |  |  |
| --- | --- | --- | --- | --- | --- | --- | --- | --- | --- |
|  | Individual<br>n (%) | Unadjusted beta<br>(95% CI) | Adjusted beta<br>(95% CI) <sup>a</sup> | Individual<br>n (%) | Unadjusted beta<br>(95% CI) | Adjusted beta<br>(95% CI) <sup>b</sup> | Individual<br>n (%) | Unadjusted beta<br>(95% CI) | Adjusted beta<br>(95% CI) <sup>b</sup> |
| **Age at separation** | 2712 |  |  | 2962 |  |  | 2687 |  |  |
| Before age 17 | 110 (4.1) | Ref. | Ref. | 131 (4.4) | Ref. | Ref. | 107 (4.0) | Ref. | Ref. |
| Age 17-19.5 | 958 (35.3) | -0.17 (-0.34, -0.01)* | -0.16 (-0.30, -0.01)* | 1068 (36.1) | -0.21 (-0.40, -0.02)* | -0.16 (-0.33, 0.02) | 950 (35.4) | -0.40 (-1.40, 0.60) | -0.15 (-1.11, 0.81) |
| Age 19.5-22 | 1143 (42.2) | -0.34 (-0.50, -0.18)* | -0.23 (-0.37, -0.08)* | 1225 (41.4) | -0.31 (-0.50, -0.12)* | -0.18 (-0.35, -0.00)* | 1130 (42.1) | -0.04 (-1.02, 0.95) | 0.14 (-0.81, 1.10) |
| After age 22 | 501 (18.5) | -0.61 (-0.79, -0.44)* | -0.43 (-0.59, -0.27)* | 538 (18.2) | -0.43 (-0.63, -0.23)* | -0.23 (-0.42, -0.05)* | 500 (18.6) | 0.65 (-0.36, 1.65) | 0.75 (-0.24, 1.74) |
| **Number of moves**<br>**before age 17** | 2755 |  |  | 3029 |  |  | 2729 |  |  |
| None | 733 (26.6) | Ref. | Ref. | 772 (25.5) | Ref. | Ref. | 725 (26.6) | Ref. | Ref. |
| Once | 730 (26.5) | 0.06 (-0.04, 0.15) | 0.05 (-0.04, 0.14) | 806 (26.6) | 0.06 (-0.04, 0.16) | 0.06 (-0.03, 0.16) | 723 (26.5) | -0.11 (-0.59, 0.37) | -0.11 (-0.59, 0.38) |
| Twice | 545 (19.8) | 0.02 (-0.08, 0.13) | 0.01 (-0.09, 0.11) | 598 (19.7) | 0.13 (0.02, 0.23)* | 0.10 (0.00, 0.20)* | 540 (19.8) | 0.55 (0.03, 1.08)* | 0.48 (-0.04, 1.01) |
| Three times or more | 747 (27.1) | 0.07 (-0.03, 0.16) | 0.01 (-0.08, 0.10) | 853 (28.2) | 0.15 (0.05, 0.24)* | 0.08 (-0.01, 0.17) | 741 (27.2) | 0.38 (-0.15, 0.91) | 0.32 (-0.21, 0.86) |
| **Total distance of moving**<br>**(per 100km) before age 17** | 2491 | 0.01 (-0.02, 0.03) | 0.01 (-0.01, 0.03) | 2731 | 0.01 (-0.02, 0.03) | 0.01 (-0.01, 0.03) | 2468 | 0.01 (-0.11, 0.13) | -0.00 (-0.11, 0.12) |

<sup>a</sup> Adjusted for sex, zygosity, smoking, work status, secondary level school, and parental education; for number of moves and total distance of moving before 17, further adjusted for separation before age 17
<sup>b</sup> Adjusted for sex, zygosity, smoking, work status, secondary level school, parental education, and age when twins provided the GBI assessment in young adulthood; for number of moves and total distance of moving before 17, further adjusted for separation before age 17
\* P<0.05

Figure 1 depicts the trajectory of the predictive marginal mean of log-transformed GBI score from age 17 to young adulthood. The three age-at-separation groups before age 22 decreased in their predictive marginal mean of log-transformed GBI scores, but the latest group seems not to change obviously. The results of the MMRM are presented in Table 3 and included 2712 twins at age 17 and 2962 twins in young adulthood. After controlling for log-transformed GBI score at age 17 and pre-specified covariates, twins who separated between ages 17 and 19.5 (beta: -0.18, 95% CI: - 0.33, -0.02), between ages 19.5 and 22 (beta: -0.20, 95% CI: -0.35, -0.04), and after age 22 (beta: -0.25, 95% CI: -0.41, -0.09) scored significantly lower on the GBI (log-transformed) in young adulthood than those who separated before age 17.

**Table 3:** Association of age at separation with GBI using MMRM

| Age at separation | Mean (SD) |  | Adjusted coefficient (95% CI) <sup>a</sup> |
| --- | --- | --- | --- |
|  | GBI at age 17<br>(individual n=2712) | GBI in young adulthood<br>(individual n=2962) | Log-transformed GBI score<br>in young adulthood |
| Before age 17 | 6.77 (5.02) | 6.20 (5.81) | Ref. |
| Age 17-19.5 | 5.92 (5.29) | 4.82 (4.90) | -0.18 (-0.33, -0.02)* |
| Age 19.5-22 | 4.91 (4.64) | 4.18 (4.38) | -0.20 (-0.35, -0.04)* |
| After age 22 | 3.66 (3.98) | 3.79 (4.50) | -0.25 (-0.41, -0.09)* |

<sup>a</sup> Adjusted for sex, zygosity, smoking, work status, secondary level school, parental education, and age when twins provided the GBI assessment in young adulthood
\* P<0.05

**Figure 1:**
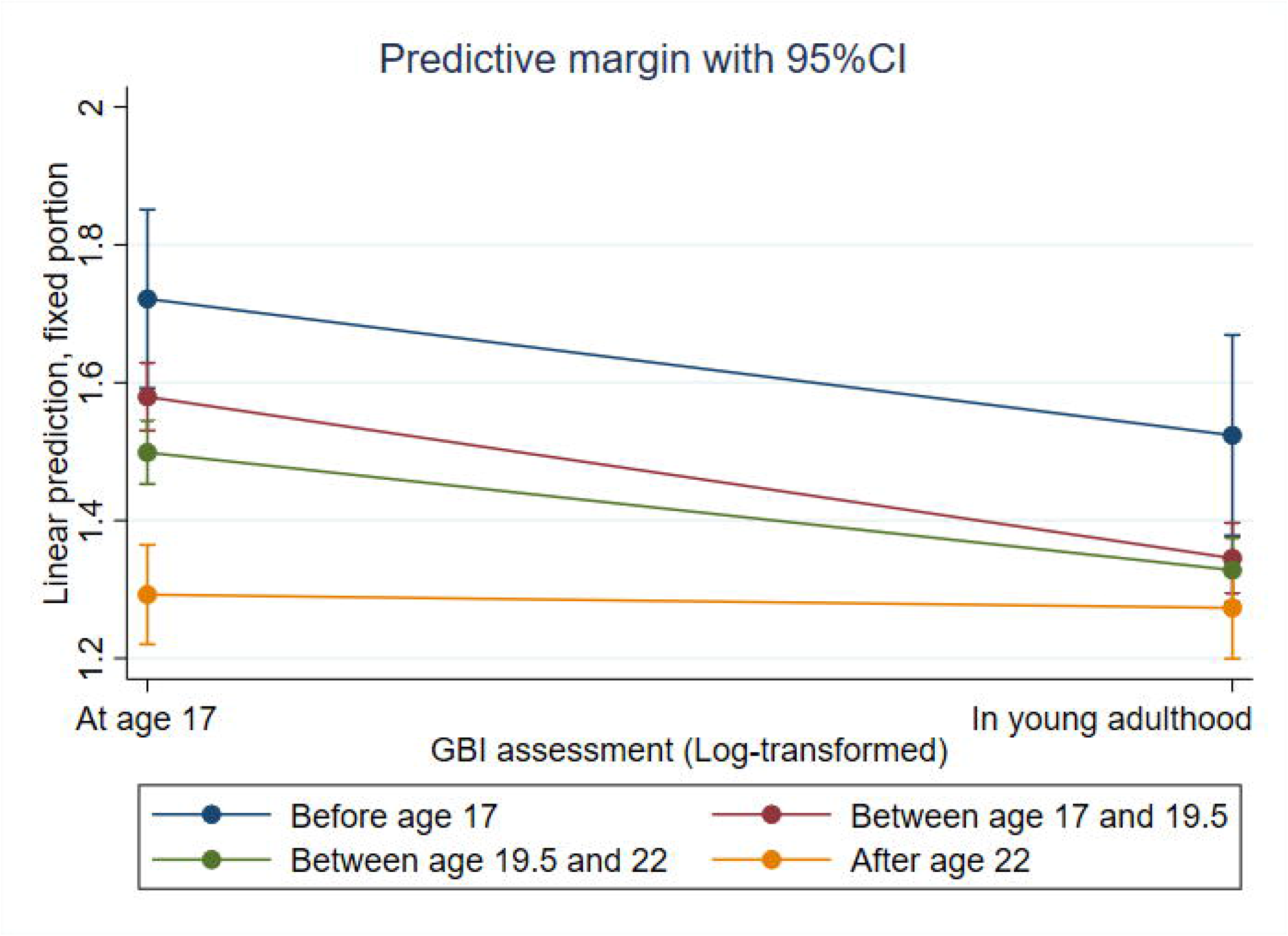
Trajectory of the predictive marginal mean of log-transformed GBI score by the age at separation

Then we performed sex stratification MMRM of age at separation (Supplemental Table 1). Regardless of the age at separation, males’ mean GBI scores were notably lower than those of females. In females, after controlling for log-transformed GBI score at age 17 and pre-specified covariates, twins who separated after age 22 had lower log-transformed GBI scores in young adulthood (beta: -0.28, 95% CI: -0.50, -0.05).

### 3.3 Association of residential mobility with GBI

In the linear regression, after adjustment for covariates, compared to never moving before age 17, moving twice increased the log-transformed GBI score at age 17 (beta: 0.10, 95% CI: 0.00, 0.20) (Table 2). However, we did not observe any significant association between the total distance of moves before age 17 and any of the GBI variables.

Supplemental Figure 1 depicts the trajectory of the predictive marginal mean of log-transformed GBI score from age 17 to young adulthood by the number of moves before age 17. The predictive marginal means decreased in all number-of moves groups and reductions were more substantial in twins who never moved or moved once than in other groups. In MMRM, after controlling for baseline values and covariates, twins who moved twice and moved three times or more scored higher GBI (log-transformed) in young adulthood than those who never moved (beta: 0.11, 95% CI: 0.02, 0.20 and beta: 0.09, 95% CI: 0.00, 0.17, respectively) (Supplemental Table 2).

### 3.4 Relationship between separation and within-pair similarity

Supplemental Table 3 presents the distribution of 1279 twin pairs over the separation status and demographic characteristics. The within-pair difference in GBI scores decreased from age 17 to age 22 overall and in almost every subgroup, except MMZ and FDZ. Cross-lagged longitudinal analyses (1058 twin pairs included) showed that within-pair difference in GBI at age 17 was significantly associated with separation status before age 22 (beta: 0.01), with a small effect size. Moreover, there were another significant associations between the within-pair differences in GBI score at ages 17 and in young adulthood (beta: 0.21) and between separation status before age 17 and age 22 (beta: 0.19) (Supplemental Figure 1).

The overall intraclass correlation coefficients of MZ and DZ pairs were 0.56 and 0.14 at age 17 and 0.52 and 0.22 in young adulthood, respectively (Supplemental Table 4). Notably, for the GBI score in young adulthood, coefficients did not differ between MZs (0.55) compared to DZs (0.53) in twin pairs who separated before age 17.

## 4. Discussion

With a total of 3055 twins from the FinnTwin12 cohort included in this study, we found that later separation in twin pairs was associated with fewer depressive symptoms in later adolescence and young adulthood. In addition, higher residential mobility led to more depressive symptoms in young adulthood. Although females had more depressive symptoms than males, there were no obvious sex differences in the association between age at separation and depressive symptoms. Furthermore, we found a small genetic influence on the separation and fairly minor and inconsistent effects of separation on the similarity of twin pairs, indicating that age at separation is not a major modifier of sibling similarity for depressive symptoms.

Leaving home is considered a hallmark of becoming an adult, in which the youth is separated from parents and siblings, lose their original familial network, and enters a completely new living environment. The majority of the age at separation of the twins in this study was between ages 17 and 22, which also implied life transition like entering university to some extent. The transition unavoidably leads to negative or positive mental health effects, which are complicated by familial contexts. Seiffe-Krenke found that the rate of family conflict was higher in adolescents who were in-time leavers than those still in the nest, and the in-time leaver pattern correlated with a worse level of psychological health [34]. However, higher individual and family income were associated with higher odds of nest-leaving [35], [36], and lower financial stress was mutually correlated with less depressive symptoms at the stage of emerging adulthood [37]. With our results, we observed that longer or more family support, indicated by an older age at separation, as well as mutual support, could help adolescents cope with the mental health burden of separation from parents and siblings (same-aged twin sibling in our case) and leaving home. Separation is regarded as a part of the social/family exposome [38]. Extended family support including more relatives has been associated with lower rates of MDD and fewer depressive symptoms [39], [40]. Longer and more support could be extrapolated to better support. Better family involvement and sibling relationships like warmth, attention, and praise predicted less MDD in adulthood and are aspects of corresponding interventions [41]–[43]. Familial cohesiveness and satisfactory social support were also shown to be helpful to depressive symptoms in lower-middle-class communities in the US [44]. Favorable family background and relationships could be protective factors to support early adulthood transition, and stable transitions reduced the depressive symptoms [45].

Residential mobility involves the constant change of the built environment and could be regarded as a part of the physical exposome. Instable residence reduces parental interaction, fragments education and growth, cuts social connections, and leads to emotional stress [46]. A solid body of evidence has been established that a strong association between higher childhood residential mobility and poorer mental health at later ages across countries [7], [9], [11]. Our findings could strengthen this association through longitudinal analysis. Solís et al. developed the biological plausibility that early negative experience increased physiological wear-and-tear, measured by allostatic load, which largely explained the health behaviors and education in later ages [47]. Nevertheless, we should be aware that the effect could be complicated by children’s emotionality, familial context, and determinates. Our null results regarding the distance of moves may be due to heterogeneous motivation, wherein a shorter distance could help to maintain the social network and longer distances could be related to work repositioning [7]. In a sample of 70 children, children with intense emotion had a surprising reduction in depressive symptoms with an increase in residential mobility, while children with non-intense emotion experienced the opposite [10]. Additionally, familial support was able to buffer the harmful effect of mobility on children’s education and career achievement [48].

There is plenty of research investigating the direction between similarity and contact, and it seems that both directions exist, but the magnitude varies by age, sex, and traits [49]–[52]. Separation is a turning point of contact, since the degree of contact inevitably decreases after separation. Based on the small effect size in the cross-lagged longitudinal analyses, the genetic influence on the separation was minimally observed. Moreover, age at separation might modify the similarity of depressive symptoms in a minor fashion. The intraclass correlation coefficients were almost always higher in MZ than in DZ at all ages at separation in our study, but the difference decreased in young adulthood and with later separation. This is consistent with increasing environmental effects with age reported in a previous study [53]. The majority of twins were separated between ages 17 and 22, which also implied more unique environmental variances. It was also corroborated by the nearly identical coefficients in the earliest separation group between MZ and DZ in young adulthood. Age at separation is capable of pinpointing the estimation of the genetic and environmental influence in future studies.

Our study was strengthened by the objective measure of the length that twins stayed in their parental home by age at separation through the national system. It could help to avoid recall bias. In addition, the two variables on residential mobility helped us quantify its cumulative and longitudinal effect on the development of twins. Furthermore, the analysis, including repeated measures, could characterize the critical phase of depressive symptom development from late adolescence to young adulthood.

There were also several limitations in this study. First, the 10-item GBI was a non-diagnostic self-report measure of depressive symptoms. However, previous studies have shown high reliability and validity of the full-scale GBI from which this shorter version is derived [54]. As we had interview-based diagnostic information on MDD on the same participants, we could show a high validity of this short GBI based on the high AUC. Second, the twin study may lead to the concern of low generalizability or representativeness. Most traits in adult twins including adaptive behavior, depression, and anxiety did not differ from singletons, which were suggested to be generalizable [55]–[57]. Third, when Finnish men are doing their military service, their residential records do not change to their service location, which may lead to information bias. Additionally, military service could possibly affect mental health [58], which suggests potential roles as a confounder or modifier. Thus, we performed the sex-stratification analysis, but there was no obvious sex-specific effect.

## 5. Conclusion

Overall, the results provided a novel and valid estimation of the effect of age at separation on depressive symptoms in later adolescence and early adulthood. It could reflect the influence of the length and amount of support from cotwin and family for twins on addressing this separation challenge and depressive symptoms. Moreover, we confirmed the association between residential mobility and mental health with longitudinal analysis. Further intervention targeting the family and living environment, could reduce the incidence of depressive symptoms.

## Data Availability

All data produced in the present study are available upon reasonable request to the authors

## Reference

[1] D. F. Santomauro et al., “Global prevalence and burden of depressive and anxiety disorders in 204 countries and territories in 2020 due to the COVID-19 pandemic,” Lancet, vol. 398, no. 10312, pp. 1700–1712, Nov. 2021, doi: 10.1016/S0140-6736(21)02143-7/ATTACHMENT/927FDFEF-CCD4-4655-AACF-4E7D54DFECF5/MMC1.PDF.

[2] E. Bromet et al., “Cross-national epidemiology of DSM-IV major depressive episode,” BMC Med., vol. 9, no. 1, p. 90, 2011, doi: 10.1186/1741-7015-9-90.

[3] A. Ferrari, “Global, regional, and national burden of 12 mental disorders in 204 countries and territories, 1990-2019: a systematic analysis for the Global Burden of Disease Study 2019,” The lancet. Psychiatry, vol. 9, no. 2, pp. 137–150, Feb. 2022, doi: 10.1016/S2215-0366(21)00395-3/ATTACHMENT/13E52F88-5D65-478F-843A-2CF574C4DFF4/MMC1.PDF.

[4] R. Mojtabai, M. Olfson, and B. Han, “National Trends in the Prevalence and Treatment of Depression in Adolescents and Young Adults,” Pediatrics, vol. 138, no. 6, p. e20161878, Dec. 2016, doi: 10.1542/peds.2016-1878.

[5] A. H. Weinberger, M. Gbedemah, A. M. Martinez, D. Nash, S. Galea, and R. D. Goodwin, “Trends in depression prevalence in the USA from 2005 to 2015: widening disparities in vulnerable groups,” Psychol. Med., vol. 48, no. 8, pp. 1308–1315, 2018, doi: DOI: 10.1017/S0033291717002781.

[6] G. Parker and K. Roy, “Adolescent Depression: A Review,” Aust. New Zeal. J. Psychiatry, vol. 35, no. 5, pp. 572–580, Oct. 2001, doi: 10.1080/0004867010060504.

[7] T. Morris, D. Manley, and C. E. Sabel, “Residential mobility: Towards progress in mobility health research,” Prog. Hum. Geogr., vol. 42, no. 1, pp. 112–133, May 2016, doi: 10.1177/0309132516649454.

[8] P. Cuijpers, F. Smit, F. Unger, Y. Stikkelbroek, M. ten Have, and R. de Graaf, “The disease burden of childhood adversities in adults: A population-based study,” Child Abuse Negl., vol. 35, no. 11, pp. 937–945, 2011, doi: https://doi.org/10.1016/j.chiabu.2011.06.005.

[9] T. Jelleyman and N. Spencer, “Residential mobility in childhood and health outcomes: a systematic review,” J. Epidemiol. Community Health, vol. 62, no. 7, pp. 584LP–592, Jul. 2008, doi: 10.1136/jech.2007.060103.

[10] Z. Stoneman, G. H. Brody, S. L. Churchill, and L. L. Winn, “Effects of Residential Instability on Head Start Children and Their Relationships with Older Siblings: Influences of Child Emotionality and Conflict between Family Caregivers,” Child Dev., vol. 70, no. 5, pp. 1246–1262, Sep. 1999, doi: https://doi.org/10.1111/1467-8624.00090.

[11] M. Simsek, R. Costa, and H. A. G. de Valk, “Childhood residential mobility and health outcomes: A meta-analysis,” Health Place, vol. 71, p. 102650, 2021, doi: https://doi.org/10.1016/j.healthplace.2021.102650.

[12] D. Silove, V. Manicavasagar, J. Curtis, and A. Blaszczynski, “Is early separation anxiety a risk factor for adult panic disorder?: A critical review,” Compr. Psychiatry, vol. 37, no. 3, pp. 167–179, 1996, doi: https://doi.org/10.1016/S0010-440X(96)90033-4.

[13] L. C. Vanderwerker, S. C. Jacobs, C. M. Parkes, and H. G. Prigerson, “An Exploration of Associations Between Separation Anxiety in Childhood and Complicated Grief in Later Life,” J. Nerv. Ment. Dis., vol. 194, no. 2, 2006, [Online]. Available: https://journals.lww.com/jonmd/Fulltext/2006/02000/An_Exploration_of_Associations_Between_Separation.8.aspx.

[14] S. G. Aschenbrand, P. C. Kendall, A. Webb, S. M. Safford, and E. Flannery-Schroeder, “Is Childhood Separation Anxiety Disorder a Predictor of Adult Panic Disorder and Agoraphobia? A Seven-Year Longitudinal Study,” J. Am. Acad. Child Adolesc. Psychiatry, vol. 42, no. 12, pp. 1478–1485, 2003, doi: https://doi.org/10.1097/00004583-200312000-00015.

[15] T. T. Swartz, M. Kim, M. Uno, J. Mortimer, and K. B. O’Brien, “Safety Nets and Scaffolds: Parental Support in the Transition to Adulthood,” J. Marriage Fam., vol. 73, no. 2, pp. 414–429, Apr. 2011, doi: https://doi.org/10.1111/j.1741-3737.2010.00815.x.

[16] T. Newman and S. Blackburn, “Transitions in the Lives of Children and Young People: Resilience Factors,” Oct. 2002.

[17] B. J. Gillespie, “Adolescent Intergenerational Relationship Dynamics and Leaving and Returning to the Parental Home,” J. Marriage Fam., vol. 82, no. 3, pp. 997–1014, Jun. 2020, doi: 10.1111/jomf.12630.

[18] L. van den Berg, M. Kalmijn, and T. Leopold, “Family Structure and Early Home Leaving: A Mediation Analysis,” Eur. J. Popul., vol. 34, no. 5, pp. 873–900, Jan. 2018, doi: 10.1007/s10680-017-9461-1.

[19] T. Egondi, C. Kabiru, D. Beguy, M. Kanyiva, and R. Jessor, “Adolescent home-leaving and the transition to adulthood: A psychosocial and behavioural study in the slums of Nairobi,” Int. J. Behav. Dev., vol. 37, no. 4, pp. 298–308, Jul. 2013, doi: 10.1177/0165025413479299.

[20] D. M. Shumaker, C. Miller, C. Ortiz, and R. Deutsch, “THE FORGOTTEN BONDS: THE ASSESSMENT AND CONTEMPLATION OF SIBLING ATTACHMENT IN DIVORCE AND PARENTAL SEPARATION,” Fam. Court Rev., vol. 49, no. 1, pp. 46–58, Jan. 2011, doi: https://doi.org/10.1111/j.1744-1617.2010.01352.x.

[21] J. Gong et al., “Sibling separation and psychological problems of double AIDS orphans in rural China – a comparison analysis,” Child. Care. Health Dev., vol. 35, no. 4, pp. 534–541, Jul. 2009, doi: https://doi.org/10.1111/j.1365-2214.2009.00969.x.

[22] K. Fortuna, I. Goldner, and A. Knafo, “Twin relationships: A comparison across monozygotic twins, dizygotic twins, and nontwin siblings in early childhood,” Fam. Sci., vol. 1, no. 3–4, pp. 205–211, Oct. 2010, doi: 10.1080/19424620.2010.569367.

[23] K. M. Mark, A. Pike, R. M. Latham, and B. R. Oliver, “Using Twins to Better Understand Sibling Relationships,” Behav. Genet., vol. 47, no. 2, pp. 202–214, Mar. 2017, doi: 10.1007/s10519-016-9825-z.

[24] M. B. Adelman and M. Siemon, “Communicating the Relational Shift: Separation among Adult Twins,” Am. J. Psychother., vol. 40, no. 1, pp. 96–109, Jan. 1986, doi: 10.1176/appi.psychotherapy.1986.40.1.96.

[25] R. J. Rose et al., “FinnTwin12 Cohort: An Updated Review,” Twin Res. Hum. Genet., vol. 22, no. 5, pp. 302–311, Oct. 2019, doi: 10.1017/thg.2019.83.

[26] Z. Wang, A. Whipp, M. Heinonen-Guzejev, and J. Kaprio, “Age at Separation of Twin Pairs in the FinnTwin12 Study,” Twin Res. Hum. Genet., pp. 1–7, 2022, doi: DOI: 10.1017/thg.2022.17.

[27] K. Kokko and L. Pulkkinen, “Unemployment and Psychological Distress: Mediator Effects,” J. Adult Dev., vol. 5, no. 4, pp. 205–217, 1998, doi: 10.1023/A:1021450208639.

[28] R. A. Depue, J. F. Slater, H. Wolfstetter-Kausch, D. Klein, E. Goplerud, and D. Farr, “A behavioral paradigm for identifying persons at risk for bipolar depressive disorder: A conceptual framework and five validation studies.,” Journal of Abnormal Psychology, vol. 90, no. 5. American Psychological Association, US, pp. 381–437, 1981, doi: 10.1037/0021-843X.90.5.381.

[29] A. Ranjit et al., “Testing the reciprocal association between smoking and depressive symptoms from adolescence to adulthood: A longitudinal twin study,” Drug Alcohol Depend., vol. 200, pp. 64–70, 2019, doi: https://doi.org/10.1016/j.drugalcdep.2019.03.012.

[30] K. Salmela-Aro et al., “Depressive Symptoms and Career-Related Goal Appraisals: Genetic and Environmental Correlations and Interactions,” Twin Res. Hum. Genet., vol. 17, no. 4, pp. 236–243, 2014, doi: DOI: 10.1017/thg.2014.33.

[31] K. K. Bucholz et al., “A new, semi-structured psychiatric interview for use in genetic linkage studies: a report on the reliability of the SSAGA.,” J. Stud. Alcohol, vol. 55, no. 2, pp. 149–158, Mar. 1994, doi: 10.15288/jsa.1994.55.149.

[32] C. Huppertz et al., “The effects of parental education on exercise behavior in childhood and youth: a study in Dutch and Finnish twins,” Scand. J. Med. Sci. Sports, vol. 27, no. 10, pp. 1143–1156, Oct. 2017, doi: https://doi.org/10.1111/sms.12727.

[33] M. A. Detry and Y. Ma, “Analyzing Repeated Measurements Using Mixed Models,” JAMA, vol. 315, no. 4, pp. 407–408, Jan. 2016, doi: 10.1001/jama.2015.19394.

[34] I. Seiffe-Krenke, “Leaving home or still in the nest? Parent-child relationships and psychological health as predictors of different leaving home patterns.,” Developmental Psychology, vol. 42, no. 5. American Psychological Association, Seiffe-Krenke, Inge: Department of Psychology, University of Mainz, Staudinger Weg 9, Mainz, Germany, Main, D-55099, pp. 864–876, 2006, doi: 10.1037/0012-1649.42.5.864.

[35] R. Avery, F. Goldscheider, and A. Speare Jr., “Feathered nest/gilded cage: Parental income and leaving home in the transition to adulthood,” Demography, vol. 29, no. 3, pp. 375–388, Aug. 1992, doi: 10.2307/2061824.

[36] C. H. Mulder and W. A. V Clark, “Leaving home and leaving the State: evidence from the United States,” Int. J. Popul. Geogr., vol. 6, no. 6, pp. 423–437, Nov. 2000, doi: https://doi.org/10.1002/1099-1220(200011/12)6:6<423::AID-IJPG199>3.0.CO;2-R.

[37] H. Cao, N. Zhou, X. Li, J. Serido, and S. Shim, “Temporal dynamics of the association between financial stress and depressive symptoms throughout the emerging adulthood,” J. Affect. Disord., vol. 282, pp. 211–218, 2021, doi: https://doi.org/10.1016/j.jad.2020.12.166.

[38] I. van Kamp et al., “Early environmental quality and life-course mental health effects: The Equal-Life project,” Environ. Epidemiol., vol. 6, no. 1, 2022, [Online]. Available: https://journals.lww.com/environepidem/Fulltext/2022/02000/Early_environmental_quality_and_life_course_mental.2.aspx.

[39] R. J. Taylor, D. H. Chae, K. D. Lincoln, and L. M. Chatters, “Extended family and friendship support networks are both protective and risk factors for major depressive disorder and depressive symptoms among African-Americans and black Caribbeans,” J. Nerv. Ment. Dis., vol. 203, no. 2, pp. 132–140, Feb. 2015, doi: 10.1097/NMD.0000000000000249.

[40] J. E. Johnson, C. Esposito-Smythers, R. Miranda, C. J. Rizzo, A. N. Justus, and G. Clum, “Gender, Social Support, and Depression in Criminal Justice–Involved Adolescents,” Int. J. Offender Ther. Comp. Criminol., vol. 55, no. 7, pp. 1096–1109, Oct. 2010, doi: 10.1177/0306624X10382637.

[41] J. T. Ehrenreich, L. C. Santucci, and C. L. Weiner, “SEPARATION ANXIETY DISORDER IN YOUTH: PHENOMENOLOGY, ASSESSMENT, AND TREATMENT,” Psicol. Conductual, vol. 16, no. 3, pp. 389–412, Jan. 2008, doi: 10.1901/jaba.2008.16-389.

[42] R. J. Waldinger, G. E. Vaillant, and E. J. Orav, “Childhood Sibling Relationships as a Predictor of Major Depression in Adulthood: A 30-Year Prospective Study,” Am. J. Psychiatry, vol. 164, no. 6, pp. 949–954, Jun. 2007, doi: 10.1176/ajp.2007.164.6.949.

[43] P. Chen and K. M. Harris, “Association of Positive Family Relationships With Mental Health Trajectories From Adolescence to Midlife,” JAMA Pediatr., vol. 173, no. 12, pp. e193336–e193336, Dec. 2019, doi: 10.1001/jamapediatrics.2019.3336.

[44] H. Z. Reinherz, G. Stewart-Berghauer, B. Pakiz, A. K. Frost, B. A. Moeykens, and W. M. Holmes, “The Relationship of Early Risk and Current Mediators to Depressive Symptomatology in Adolescence,” J. Am. Acad. Child Adolesc. Psychiatry, vol. 28, no. 6, pp. 942–947, 1989, doi: https://doi.org/10.1097/00004583-198911000-00021.

[45] A. Booth, E. Rustenbach, and S. McHale, “Early Family Transitions and Depressive Symptom Changes From Adolescence to Early Adulthood,” J. Marriage Fam., vol. 70, no. 1, pp. 3–14, Feb. 2008, doi: https://doi.org/10.1111/j.1741-3737.2007.00457.x.

[46] E. Baker, N. T. A. Pham, L. Daniel, and R. Bentley, “How Does Household Residential Instability Influence Child Health Outcomes? A Quantile Analysis,” Int. J. Environ. Res. Public Health, vol. 16, no. 21, p. 4189, Oct. 2019, doi: 10.3390/ijerph16214189.

[47] B. S. Cristina et al., “Adverse childhood experiences and physiological wear-and-tear in midlife: Findings from the 1958 British birth cohort,” Proc. Natl. Acad. Sci., vol. 112, no. 7, pp. E738–E746, Feb. 2015, doi: 10.1073/pnas.1417325112.

[48] J. Hagan, R. MacMillan, and B. Wheaton, “New Kid in Town: Social Capital and the Life Course Effects of Family Migration on Children,” Am. Sociol. Rev., vol. 61, no. 3, pp. 368–385, Jun. 1996, doi: 10.2307/2096354.

[49] S. F. Posner, L. Baker, A. Heath, and N. G. Martin, “Social contact, social attitudes, and twin similarity,” Behav. Genet., vol. 26, no. 2, pp. 123–133, 1996, doi: 10.1007/BF02359890.

[50] J. Kaprio, M. Koskenvuo, and R. J. Rose, “Change in cohabitation and intrapair similarity of monozygotic (MZ) cotwins for alcohol use, extraversion, and neuroticism,” Behav. Genet., vol. 20, no. 2, pp. 265–276, 1990, doi: 10.1007/BF01067794.

[51] D. T. Lykken, M. McGue, T. J. Bouchard, and A. Tellegen, “Does contact lead to similarity or similarity to contact?,” Behav. Genet., vol. 20, no. 5, pp. 547–561, 1990, doi: 10.1007/BF01065871.

[52] R. J. Rose, J. Kaprio, C. J. Williams, R. Viken, and K. Obremski, “Social contact and sibling similarity: Facts, issues, and red herrings,” Behav. Genet., vol. 20, no. 6, pp. 763–778, 1990, doi: 10.1007/BF01065919.

[53] M. G. Nivard et al., “Stability in symptoms of anxiety and depression as a function of genotype and environment: a longitudinal twin study from ages 3 to 63 years,” Psychol. Med., vol. 45, no. 5, pp. 1039–1049, 2015, doi: DOI: 10.1017/S003329171400213X.

[54] L. L. Pendergast, E. A. Youngstrom, C. Brown, D. Jensen, L. Y. Abramson, and L. B. Alloy, “Structural invariance of General Behavior Inventory (GBI) scores in Black and White young adults,” Psychol. Assess., vol. 27, no. 1, pp. 21–30, Mar. 2015, doi: 10.1037/pas0000020.

[55] M. A. Munn-Chernoff, K. M. von Ranson, K. M. Culbert, C. L. Larson, S. A. Burt, and K. L. Klump, “An examination of the representativeness assumption for twin studies of eating pathology and internalizing symptoms,” Behav. Genet., vol. 43, no. 5, pp. 427–435, Sep. 2013, doi: 10.1007/s10519-013-9603-0.

[56] L. Pulkkinen, I. Vaalamo, R. Hietala, J. Kaprio, and R. J. Rose, “Peer Reports of Adaptive Behavior in Twins and Singletons: Is Twinship a Risk or an Advantage?,” Twin Res., vol. 6, no. 2, pp. 106–118, 2003, doi: DOI: 10.1375/twin.6.2.106.

[57] G. Willemsen, V. Odintsova, E. de Geus, and D. I. Boomsma, “Twin-Singleton Comparisons Across Multiple Domains of Life BT - Twin and Higher-order Pregnancies,” A. Khalil, L. Lewi, and E. Lopriore, Eds. Cham: Springer International Publishing, 2021, pp. 51–71.

[58] K. Appelqvist-Schmidlechner, M. Upanne, M. Henriksson, K. Parkkola, and E. Stengå Rd, “Young men exempted from compulsory military or civil service in Finland-A group of men in need of psychosocial support?,” Scand. J. Public Health, vol. 38, pp. 168–176, 2010, doi: 10.1177/1403494809357103.

