## Supplementary Table 1-4 and Figure 1-2 for "Age at separation, residential mobility, and depressive symptoms among twins in late adolescence and young adulthood: a FinnTwin12 cohort study"

**Zhiyang Wang et al. – Online supplementary material**

### Content of Supplementary Material

Supplemental Table 1: Sex-stratified association of age at separation with GBI using MMRM

Supplemental Table 2: Association of number of moves before age 17 with GBI using MMRM

Supplemental Table 3: Separation status, demographic characteristics, and within-pair differences in GBI score for twin pairs

Supplemental Table 4: Intraclass correlation coefficients (r) among MZ and DZ twin pairs

Supplemental Figure 1: Trajectory of the predictive marginal mean of log-transformed GBI score by the number of moves

Supplemental Figure 2: Cross-lagged path model for within-pair differences in GBI score and separation status (1279 twin pairs). Numbers over the lines indicate the regression coefficients.

Supplemental Table 1: Sex-stratified association of age at separation with GBI using MMRM

| Age at separation | Mean (SD) | | Adjusted coefficient (95% CI) [a] |
|---|---|---|---|
| | GBI at age 17 | GBI in young adulthood | Log-transformed GBI score in young adulthood |
| *In males (1153 individual twins at age 17 and 1287 individual twins in young adulthood)* | | | |
| Before age 17 | 4.89 (4.04) | 4.79 (4.96) | Ref. |
| Age 17-19.5 | 3.94 (4.03) | 3.86 (4.63) | -0.20 (-0.42, 0.03) |
| Age 19.5-22 | 3.80 (3.99) | 3.41 (3.86) | -0.21 (-0.43, 0.01) |
| After age 22 | 2.87 (3.47) | 3.48 (4.55) | -0.23 (-0.46, 0.01) |
| *In females (1559 individual twins at age 17 and 1675 individual twins in young adulthood)* | | | |
| Before age 17 | 8.03 (5.24) | 7.28 (6.21) | Ref. |
| Age 17-19.5 | 6.90 (5.56) | 5.33 (4.97) | -0.15 (-0.36, 0.05) |
| Age 19.5-22 | 5.86 (4.93) | 4.86 (4.69) | -0.17 (-0.38, 0.04) |
| After age 22 | 4.53 (4.32) | 4.15 (4.44) | -0.28 (-0.50, -0.05)* |

[a] Adjusted for zygosity, smoking, secondary level school, work, parental education, and age when twins provided the GBI assessment in young adulthood

* P<0.05

Supplemental Table 2: Association of number of moves before age 17 with GBI using MMRM

| Number of moves before age 17 | Mean (SD) | | Adjusted coefficient (95%CI) [a] |
|---|---|---|---|
| | GBI score at age 17 (individual n=2755) | GBI score in young adulthood (individual n=3029) | Log-transformed GBI score in young adulthood |
| None | 4.86 (4.61) | 3.95 (4.20) | Ref. |
| Once | 5.20 (5.00) | 4.25 (4.53) | 0.06 (-0.02, 0.14) |
| Twice | 4.97 (4.70) | 4.73 (5.10) | 0.11 (0.02, 0.20)* |
| Three times or more | 5.31 (5.06) | 4.78 (4.89) | 0.09 (0.00, 0.17)* |

[a] Adjusted for sex, zygosity, smoking, secondary level school, work, parental education, separation before age 17, and age when twins provided the GBI assessment in young adulthood

Supplemental Table 3: Separation status, demographic characteristics, and within-pair differences in GBI score for twin pairs

| Characteristics | Twin pair n (%) | Mean (SD) Within-pair difference of GBI score | |
|---|---|---|---|
| | | At age 17 [c] | In young adulthood [d] |
| **Overall** | 1279 | 3.93 (4.10) | 3.65 (3.85) |
| **Separation before age 17** [a] | | | |
| Yes | 52 (4.1) | 4.43 (3.78) | 4.16 (4.10) |
| No | 1207 (95.9) | 3.94 (4.13) | 3.65 (3.86) |
| **Separation before age 22** [a] | | | |
| Yes | 1031 (81.9) | 4.16 (4.20) | 3.77 (3.89) |
| No | 228 (18.1) | 3.05 (3.60) | 3.25 (3.78) |
| **Sex and zygosity combination** [b] | | | |
| MMZ | 176 (14.6) | 2.32 (2.70) | 2.63 (3.33) |
| FMZ | 262 (21.8) | 3.07 (2.72) | 3.04 (2.95) |
| MDZ | 178 (14.8) | 3.87 (3.87) | 3.51 (3.89) |
| FDZ | 227 (18.9) | 4.41 (4.17) | 4.44 (4.34) |
| OSDZ | 360 (29.9) | 5.01 (4.93) | 4.19 (4.02) |
| **Parental education** | | | |
| Limited | 717 (56.1) | 4.02 (4.19) | 3.66 (3.72) |
| Intermediate | 291 (22.8) | 3.76 (3.82) | 3.60 (3.80) |
| High | 271 (21.2) | 3.91 (4.17) | 3.69 (4.26) |

[a] 20 twin pairs were missing due to age at separation

[b] 76 twin pairs were missing due to unknown zygosity

[c] 125 twin pairs were missing because at least one of the cotwins in pairs did not provide GBI assessment at age 17

[d] 22 twin pairs were missing because at least one of the cotwins in pairs did not provide GBI assessment in young adulthood

Supplemental Table 4: Intraclass correlation coefficients (r) among MZ and DZ twin pairs

| Intraclass correlation (Total 1279 twin pair) | GBI score at age 17 [a] | | GBI score in young adulthood [b] | |
|---|---|---|---|---|
| | MZ twin pair r (n twin pairs) | DZ twin pair r (n twin pairs) | MZ twin pair r (n twin pairs) | DZ twin pair r (n twin pairs) |
| **Overall** | 0.56 (431) | 0.14 (678) | 0.52 (466) | 0.22 (737) |
| **Age at separation [c]** | | | | |
| Before age 17 | 0.63 (16) | 0.14 (23) | 0.55 (20) | 0.53 (28) |
| Age 17-19.5 | 0.63 (137) | 0.14 (255) | 0.58 (153) | 0.14 (278) |
| Age 19.5-22 | 0.46 (179) | 0.06 (279) | 0.52 (187) | 0.19 (301) |
| After age 22 | 0.50 (86) | 0.16 (118) | 0.40 (92) | 0.33 (127) |

[a] 170 twin pairs were missing due to unknown zygosity or no GBI score in at least one cotwin at age 17

[b] 76 twin pairs were missing due to unknown zygosity or no GBI score in at least one cotwin in young adulthood

[c] Additional 16 and 17 twin pairs were missing due to age at separation for GBI score at age 17 and in young adulthood, respectively

Supplemental Figure 1: Trajectory of the predictive marginal mean of log-transformed GBI score by the number of moves

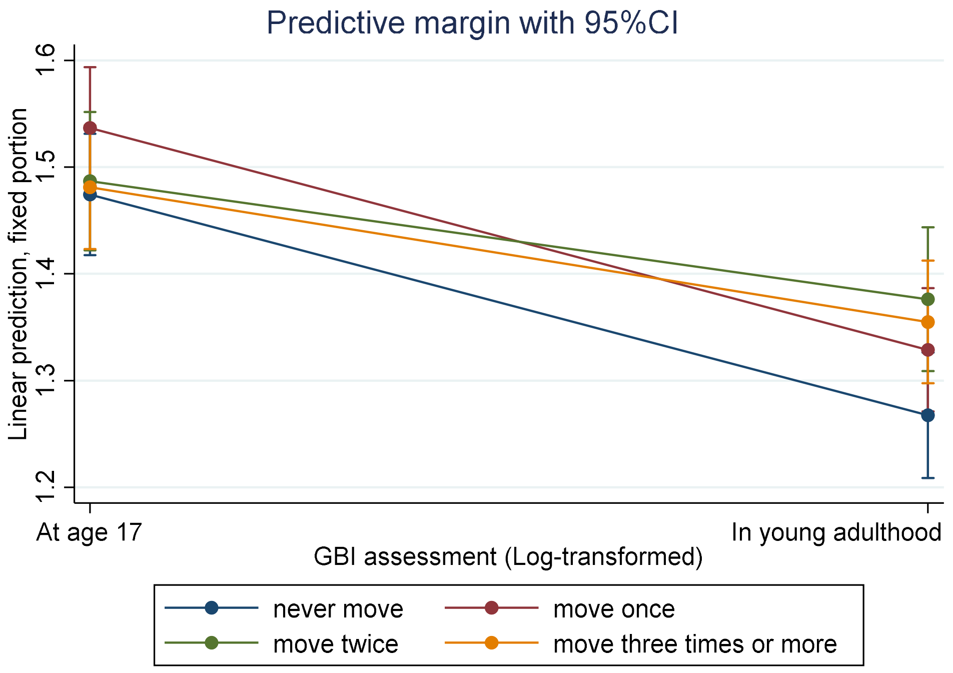

Supplemental Figure 2: Cross-lagged path model for within-pair differences in GBI score and separation status (1279 twin pairs). Numbers over the lines indicate the regression coefficients (95% CI).

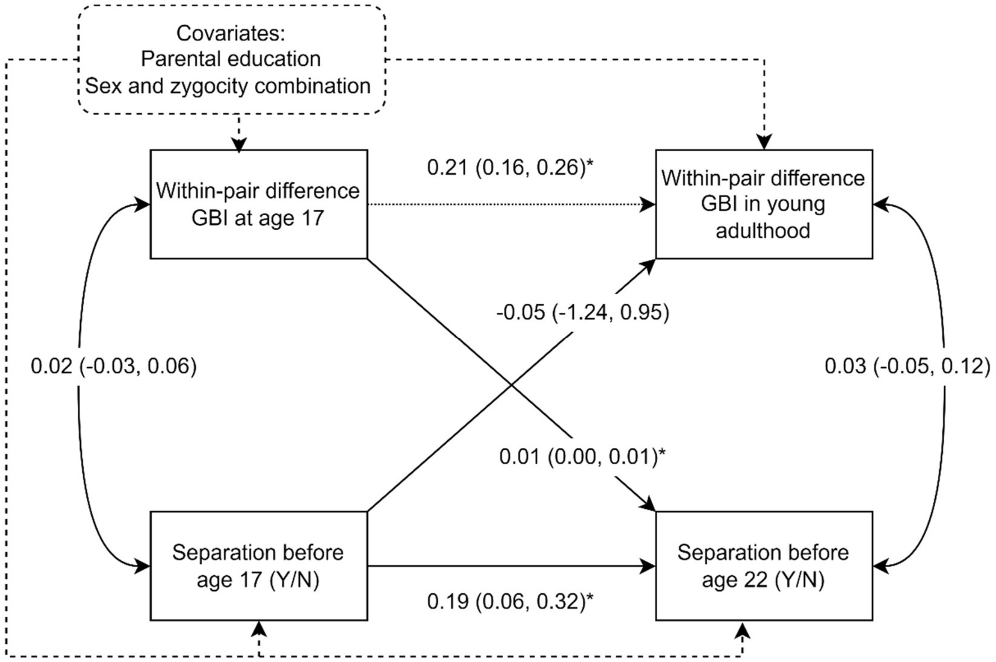


* P<0.05

Note: 221 twin pairs, whose zygosity were unknown or who were missing information on separation status and within-pair differences in GBI score at both stages, were excluded
